# Safety and efficacy of long-acting insulins (degludec and glargine) among type 2 diabetic Asian Population: A Systematic Review and Meta-Analysis

**DOI:** 10.1101/2021.01.27.21250620

**Authors:** R Kant, P Yadav, M Garg, Y Bahurupi, B Kumar

## Abstract

**Background:** According to IDF Diabetes Atlas 2019, globally, 463 million people live with Diabetes mellitus. Out of that, 88 million people are in South East Asia. By 2045, it is expected to increase by 51% globally and 74% in South East Asia. Global variation in susceptibility to diabetes, insulin sensitivity, and regimen intensity due to race and ethnic differences pose a challenge regarding the optimal choice of second-line therapy for clinicians. Asian populations are at higher risk of developing diabetes mellitus than the European population. The current study was carried out to see the relative efficacy of currently available long-acting insulins in reducing blood sugar, HbA1c and the occurrence of hypoglycemia as a complication associated with them.

**Methods:** A systematic literature search was done using various search engines (PubMed, Cochrane, Google Scholar, Scopus, and Embase) and included published RCTs in English before December 2019. Further, a manual search was performed by screening the reference list of the identified articles.

**Results:** We included four RCTs with 534 participants (349 in the insulin degludec group and 185 in the insulin glargine group) with T2DM. Results show that both insulin glargine and degludec are equally efficacious in reducing fasting blood glucose and HbA1c. However, insulin glargine was associated with lower risks of hypoglycemia.

**Conclusions:** Insulin glargine and degludec are comparable in achieving glycemic control with fewer hypoglycemic episodes in insulin glargine treated group.

## INTRODUCTION

Type 2 Diabetes Mellitus is a chronic non-communicable disease characterized by progressive B- cell dysfunction. ^[1]^ It occurs due to hyperglycemia and insulin resistance leading to increased insulin demands of tissue that culminates into over functioning of B-cells and ultimately leading to B-cell failure. ^[2]^ According to IDF Diabetes Atlas 2019, globally, 463 million people live with Diabetes mellitus, and out of that, 88 million people are from South East Asia region (SEAR) only. By 2045, it is expected to increase to 51% globally and 74% in SEAR.^[3]^ Global variation in susceptibility to diabetes, insulin sensitivity, and regimen intensity due to race and ethnic differences pose a challenge regarding the optimal choice of second-line therapy for clinicians. ^[4-6]^ Asian population is not only at higher risk of developing type 2 diabetes mellitus than European but also develop diabetes at lower BMI. ^[7- 9]^ In South-East Asian Region, amongst people with diabetes, 90% population has type 2 diabetes mellitus, which is preventable ^[10].^ Glycemic control is the only way to reduce microvascular, & macrovascular complications of DM, which can be attained by effective pharmacotherapeutic agents.^[11]^ Currently, available options for treatment include oral antidiabetic drugs and insulin Oral antidiabetic agents are often limited in their efficacy to reduce HbA1c beyond 1-2%.^[12]^ Insulin is the only drug that can reduce HbA1c to exceptionally lower levels and maintain it near normal. However, it’s use is often associated with episodes of hypoglycemia, which limits its use.^[13]^ Newer long-acting insulins is are better stable and efficacious to conventional insulin.^[14]^ There are conflicting results in earlier studies like a meta-analysis by Su W et al., found Glargine was to be superior to degludec. ^[15]^ Contrary to this, meta-analysis by Zhou W et al., found degludec much better than glargine.^[16]^ In view of the conflicting data regarding efficacy, it is necessary to compare and prioritize the treatment so far as long-acting insulin use is concerned. It was altogether more important to undertake studies in Asian population as earlier meta-analysis hardly included this population. This study aimed to evaluate the safety and efficacy of currently available long-acting insulin (insulin degludec and glargine) in type 2 diabetes mellitus Asian patients.

### Aim

To compare the efficacy and safety of long-acting insulins (degludec and glargine) in type 2 diabetic Asian population.

### Objectives

1. Efficacy assessment: - change in HbA1c and fasting plasma glucose.
2. Safety assessment: - episodes (>1) of documented hypoglycemia (< 70 mg/dl).

### Data Sources and Search strategy

The literature search was done as per the PRISMA protocol (preferential reporting items for systematic reviews and meta-analysis) ^[17]^ (figure 1) and PICO format.

**Figure 1.**
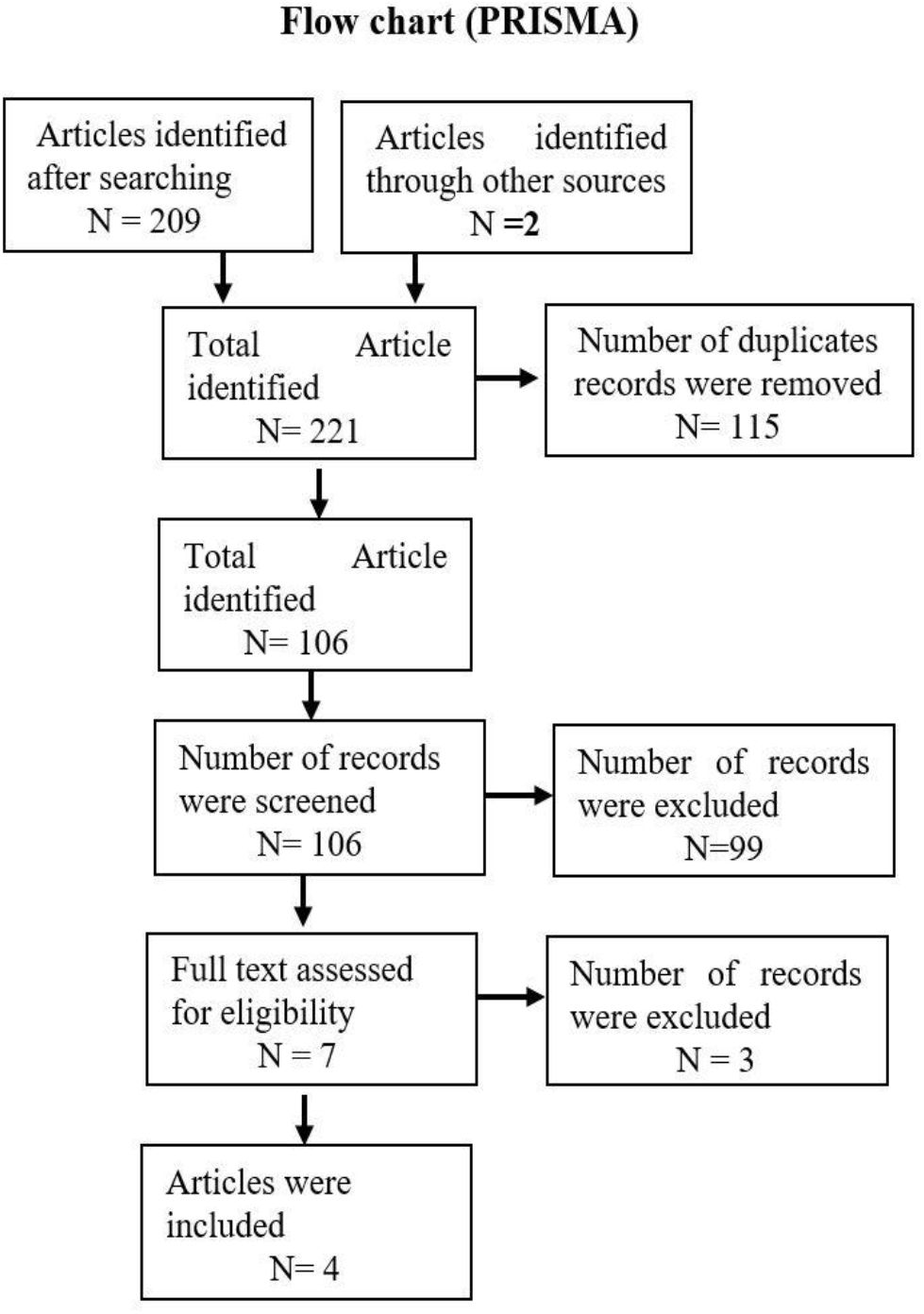
PRISMA flow chart.

Our search strategy includes all relevant published RCTs before December 2019 from PubMed, Cochrane, Google Scholar, and Embase. Articles in English format and having an Asian population were included. Keywords which were used for search (“insulin Degludec” OR “degludec” OR “IDeg”) AND (“insulin glargine” OR “insulin” “glargine” OR “IGlar”) AND (“randomized” OR “randomly”) AND (“diabetes” OR “diabetes mellitus” OR “type 2 diabetes mellitus, OR “type 2 diabetes”) AND (“Asians”). Further, a manual search was performed by screening the reference list of the identified articles.

### Inclusion criteria

1. Patient population: all studies enrolling Asian patients
2. Intervention: insulin degludec
3. Comparison: insulin glargine.
4. Outcome: safety was assessed with a risk of hypoglycemia, and efficacy was assessed by improvement or change in the HbA1c or fasting plasma glucose.
5. Study design: RCT and parallel and cross-over studies

### Exclusion criteria

1. Articles on animal studies and studies conducted outside Asia
2. Review articles, observational, quasi-experimental studies, and case series
3. Studies in which duration of intervention was less than 12 weeks.

### Data extraction

Two reviewers independently screened the studies for eligibility and data extraction. Any disagreements and discrepancies were resolved by consensus with a third reviewer. The following data from eligible studies were extracted without modification of original data onto pre-existing data collection forms: study location, year of publication, sample size, types of sample selection and allocation to two groups (intervention and comparator or control group), methods, types of administration of intervention and control, and primary outcomes. Authors of selected articles have been contacted for the required data for analysis. The search file was imported to the Zotero library. After removing duplicate items, a freely available Rayyan (
https://rayyan.qcri.org), free web-based software, was used to exclude studies.^[18]^

### Evaluation of the risk of bias (quality) assessment

The risk of bias has been evaluated through the Cochrane Risk of Bias Assessment Tool. Two reviewers assessed the risk of bias independently, and the third reviewer was consulted in case of any disagreement at any point. The risk of bias assessment has been calculated undertaking blinding (masking) of participants, researcher and outcomes, allocation concealment, randomization details, data outcome, and data reporting.^[19]^ Quality assessments have been done with GRADEpro GDT software. Relevant file has been imported from Rev Man software to “Summary of Findings” table GRADE Profiler to create a “Summary of Findings” table. ^[20]^ A summary of the intervention effect and a measure of the quality of evidence was noted in the table.

### Data analysis

Four RCTs that met the eligibility criteria were enrolled.^[21-24]^ Onishi et al. considered the confirmed hypoglycemia as plasma glucose level <3.1 mmol/L or 55.86 mg/dl and found lower hypoglycemia rates in degludec treated patients as opposed to the glargine group.^[23]^ These findings are reinforced by Aso Y et al., with a similar improvement in glycemic control in both groups but a low risk of hypoglycemia with degludec, considering plasma glucose level as < 70 mg/dl for confirmed hypoglycemia.^[21]^ In contrast, Kawaguchi Y et al. ^[22]^ and Yamabe M et al. ^[24]^ considered the confirmed hypoglycemia as plasma glucose level <70 mg/dl and observed that insulin glargine and insulin degludec are equivalent long-acting insulin analogs and insulin glargine had lesser episodes of hypoglycemia. They noted the mean percentage of time in the target glucose range (70– 179 mg/dL) and considered hypoglycemia as <70 mg/dL, using flash glucose monitoring every seven days. Authors of two studies Kawaguchi Yet al, 2019 ^[22]^, and Yamabe M et al. 2019 ^[24]^ have been contacted for the number of events with hypoglycemia episodes and post-intervention data for change in HbA1c (mean and standard deviation) for the inclusion of studies in the meta-analysis, but due to unavailability of data, these two studies could not be included in final assessment.

Relevant articles were tabulated on a Forest plot, and results were displayed based on the graph. The I^2^ statistic was used to test statistical heterogeneity, which may arise due to inter-trial variability, with>75% values representing important heterogeneity with a fixed-effects model (95% confidence interval, p-value < 0.05). For calculating efficacy, the mean difference between insulin degludec and glargine group was calculated to show differential mean changes in HbA1c and fasting plasma glucose (FPG) levels. For calculating safety between insulin degludec and glargine group, the risk ratio (RR) was calculated. We have not observed significant heterogeneity for the incidence of hypoglycemia and glycemic control. Sensitivity analysis was performed for the outcome of control of fasting plasma glucose, although we observed similar heterogeneity with random-effect model. Subgroup analysis could not be possible due to limited number of studies.

## Results

**-** Analysis was done using the Review Manager 5.4 software. Finally, four RCTs with a total of 534 participants (349 in the insulin degludec group and 185 in the insulin glargine group) with type 2 diabetes mellitus were eligible for inclusion in this Meta-analysis. Characteristics of participants in each trial have been described **in table 1**.

**Table 1.** shows characteristics of participants in each trial.

**Table1 shows characteristics of participants in each trial**
| Author and year of the study | Setting | Population | Number of Participants | Duration of Trial | Mean Age of Participants | Duration of Diabetes | Gender Distribution |
| --- | --- | --- | --- | --- | --- | --- | --- |
| Aso Y et al., 2017 <sup>[21]</sup> | A Single-Center Study | Japan | 45 participants [33 in insulin degludec group and 12 in glargine group] | 24 weeks | 64.0±13.6 in insulin degludec and 64.7±15.7 in insulin glargine | 10 years in degludec and 13 years in insulin glargine | Female participants 55% |
| Kawaguchi Yet al, 2019 <sup>[22]</sup> | Single Center Study | Japan | 30 patients [15 patients in each group] | 6 months | 69.5 ± 11.3 years | 18.3 ± 11.3 years | Male participants (60%) |
| Onishi Y et al., 2013 <sup>[23]</sup> | A Multi-center Study | Hong Kong, Japan, Malaysia, South Korea, Taiwan, and Thailand) | 435 participants [289 in insulin degludec group and 146 in glargine group] | 26 weeks | 58.6 years | 11.6 Years | Male participants (53.56%) |
| Yamabe M et al., 2019 <sup>[24]</sup> | Single Center Study | Japan | 24 participants [12 in each group] | 5 months | 70.7 ± 7.6 (years) | 14.0 ± 9.3 (years) | Male (50%) |

The summary of findings with the grade of evidence has been shown in **table 2**. This table has displayed the certainty of evidence and grade as high, moderate, and low for each outcome variable.

**Table 2.**
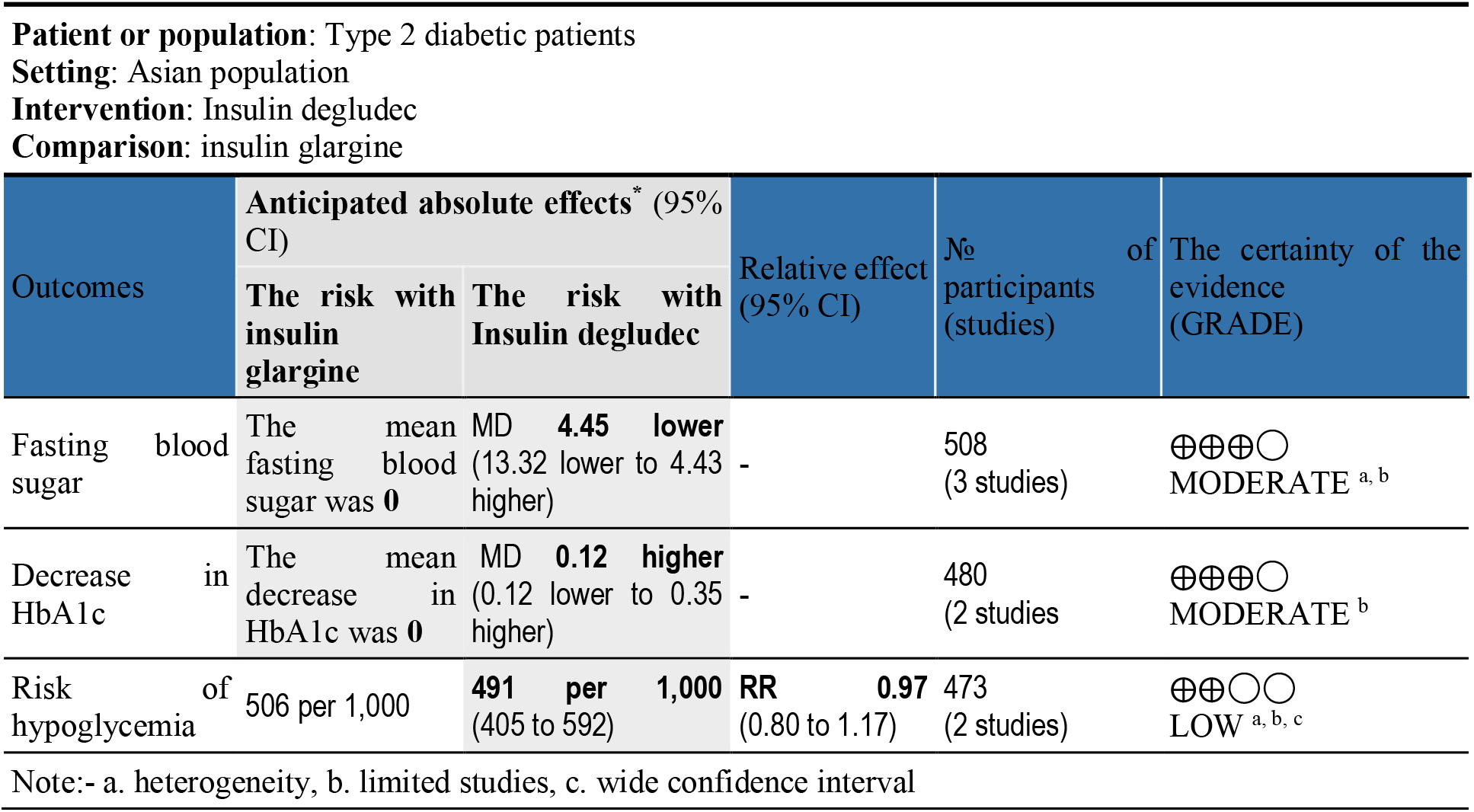
GRADEpro Summary of findings.

Figure 2 (A) forest plot for hypoglycemia risk shows RR = 0.9684, suggesting that hypoglycemia is less with glargine, CI of RR (0.8003- 1.1717) with I^2^=30%, which shows mild heterogeneity among studies.

**Figure 2.**
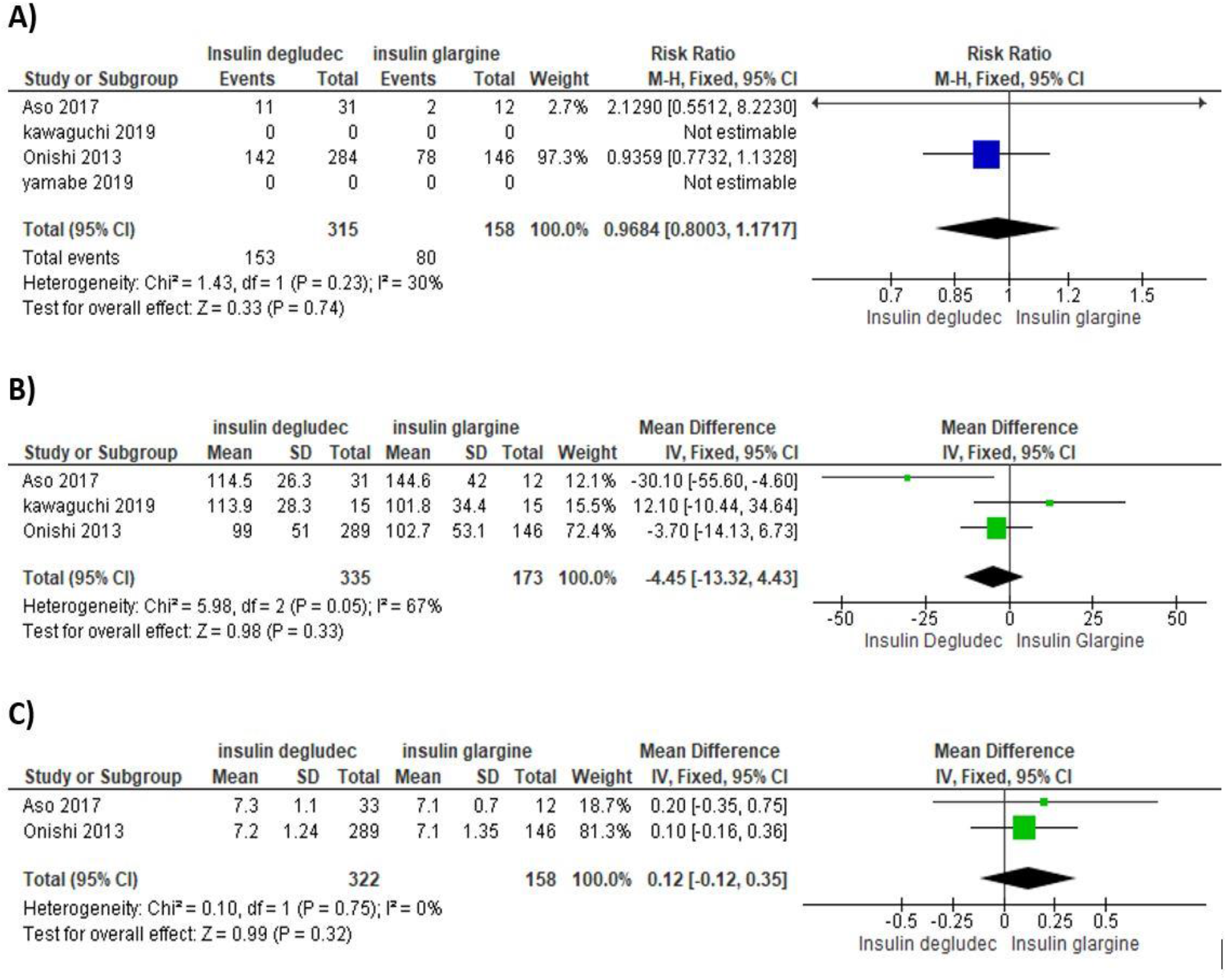
Forest plot shows the comparison of insulin degludec and glargine. A) Incidence of hypoglycemia B) Control of fasting plasma glucose, C) Decrease in HbAlc

Figure 2 (B) forest plot states no statistically significant difference in fasting plasma glucose between insulin degludec and insulin glargine [mean difference is -4.45, confidence interval - 13.32- 4.43] with I^2^=67%, p-value =0.02, which shows heterogeneity among studies. Figure 2 (C) forest plot shows no statistically significant difference in HbA1c between insulin degludec and insulin glargine [mean difference is 0.12, confidence interval -0.12-0.35] with I^2^=0%.

The risk of bias graph and summary has been displayed in **Figure 3**, which included random sequence generation, allocation concealment, detection bias, attrition bias, reporting bias, and others in each study. Funnel plots have not been created due to limited studies in the analysis.

**Figure 3.**
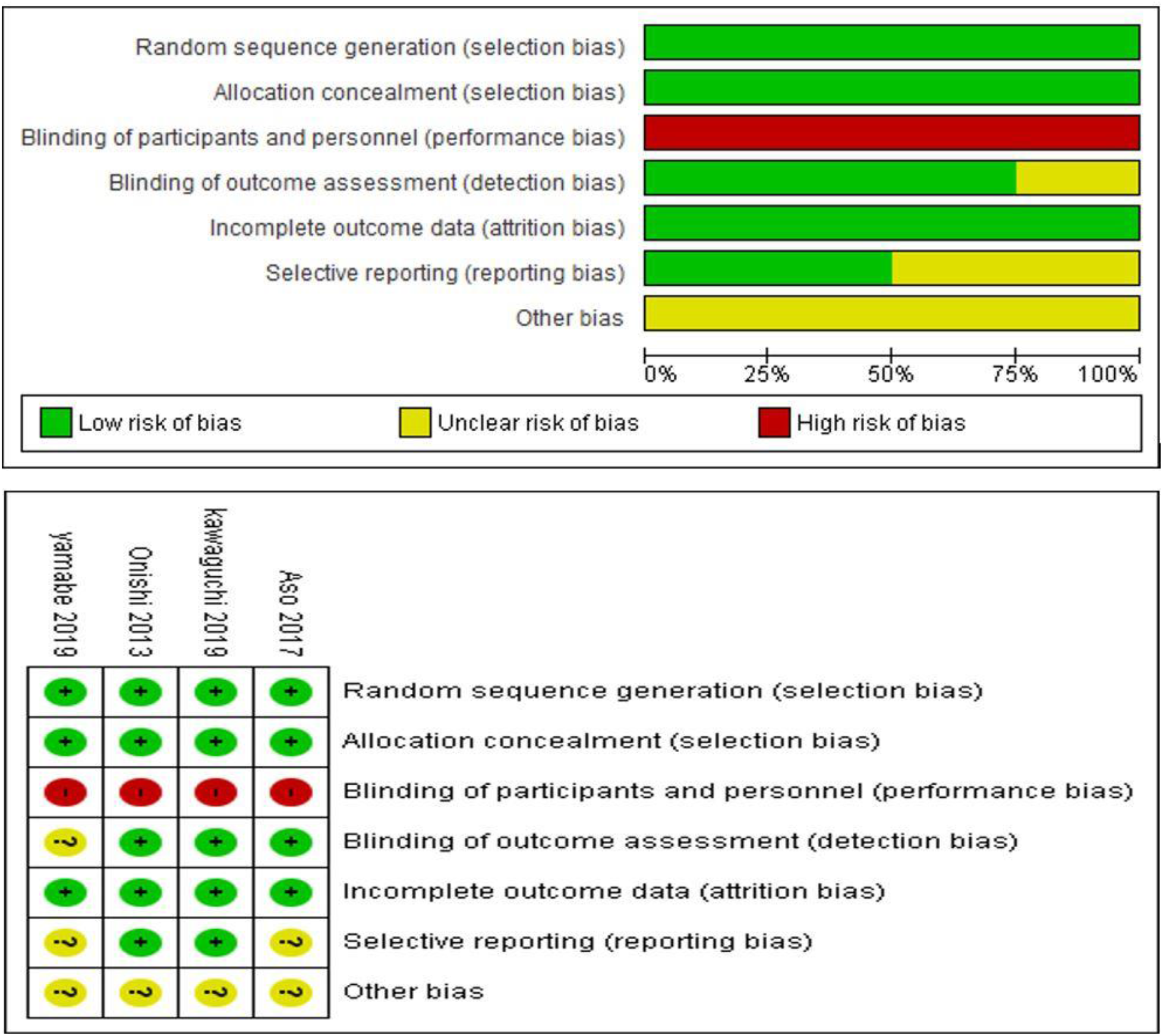
Risk of bias graph and summary.

## Discussion

Optimum glycemic control in type 2 diabetes patients represents a challenge even with advanced pharmacologic treatment.^[11]^ Evidence suggested that insulin therapy has been convincingly found to be superior in attaining target glycemic control over oral anti-diabetic drugs.^[12]^ However, its use is accompanied by hypoglycemia episodes; long-acting insulin therapy is a safe and effective way of treating diabetes mellitus.^[13]^ In the present systematic review and meta-analysis, we compared the safety and efficacy of currently available long-acting insulin (insulin degludec and glargine) in type 2 diabetes mellitus, Asian patients. Total four high-quality RCTs were included to evaluate outcome measures. In the present study, both insulins were equally effective in reducing fasting plasma glucose and HbA1c. We have noted no statistically significant mean difference in fasting plasma glucose [mean difference is -4.45, confidence interval -13.32- 4.43] and HbA1c reduction [mean difference is 0.12, confidence interval -0.12-0.35] between both groups. For comparing safety between the groups, the risk of hypoglycemia was low in glargine group RR = 0.9684, suggestive that risk of hypoglycemia is less with glargine, CI of RR (0.8003- 1.1717) with I^2^=30%. A meta-analysis by Heller S et al. showed a lower rate of hypoglycemia in the degludec group. ^[26]^ Findings of this study are supported by Onishi et al. and Pan C et al. both these investigators considered the confirmed hypoglycemia as plasma glucose level <3.1 mmol/L or 55.86 mg/dl and did find lower rates of hypoglycemia in degludec treated patients as opposed to glargine group.^[23, 25]^ These findings are reinforced by Aso Y et al. also with a similar improvement in glycemic control in both groups but a low risk of hypoglycemia with degludec, considering higher plasma glucose level (< 70 mg/dl) for confirmed hypoglycemia.^[21]^ Kawaguchi Y et al. ^[22]^ and Yamabe M et al. ^[24]^ also considered the confirmed hypoglycemia as plasma glucose level <70 mg/dl and observed that insulin glargine and insulin degludec are equivalent long-acting insulin analogs and insulin glargine has lesser episodes of hypoglycemia. Meta-analysis by Ratner RE et al. also concluded that similar improvements in glycemic control could be achieved along with fewer hypoglycemic episodes with insulin glargine.^[27]^ Findings of Ratner RE et al. and Rodbard HW et al. were conflicting, as later concluded that insulin degludec had fewer hypoglycemic events than insulin glargine ^[28],^ although clinical efficacy was similar. Even Russell-Jones D et al. reported a higher clinical efficacy and safety of insulin degludec than insulin glargine among patients with type 2 diabetes.^[29]^ Gough SC et al. in a randomized controlled trial among T2DM patients, also concluded that insulin degludec improved glycemic control similar to insulin glargine with a low risk of hypoglycemia.^[30]^ It is prudent from the studies that most researchers found a lower hypoglycaemic profile in patients treated with insulin degludec. These results could be partially explained by the cut-off value of plasma glucose to define hypoglycemia as a value of 55.86 mg/dl or less than the plasma sugar of 70mg/dl is expected to exclude a fair number of patients with hypoglycemia. Our findings are contrary to most available literature as in the current meta-analysis, we found that patients with insulin glargine were associated with lower rates of hypoglycemia. However, both insulins had equal propensity to achieve short term and long term glycemic goals. This systematic review and meta-analysis are novel work as there were only a few studies conducted among the Asian population to compare the efficacy and safety of currently available long-acting insulin (insulin degludec and glargine) in type 2 diabetes mellitus patients.

Limitations: - Studies included in this meta-analysis were open-label design, and results represents heterogeneity.

## Conclusions

This meta-analysis concluded similar efficacy of currently available long-acting insulin (insulin degludec and glargine) in type 2 diabetes mellitus patients in Asian region, with lower hypoglycemia episodes with insulin glargine.

## Data Availability

Data included in meta-analysis have been presented in the manuscript.

## Acknowledgment

None

